# Atrial Fibrillation Screening in High-Risk Patients Using a Single Lead ECG Device: A Tertiary Hospital Experience

**DOI:** 10.1101/2023.08.09.23293906

**Authors:** Tesfamariam Betemariam Aklilu, Meskerem Aleka Kebede, Duffera Mekonnen, Yidnekachew Birhan Asrat, Adane Sikamo Petros, Desalew Mekonnen

**Author notes:** Address for Correspondence: Tesfamariam Betemariam Aklilu, Affiliation: Addis Ababa University, Address: Addis Ababa, Ethiopia.

## Abstract

**Background:** Atrial Fibrillation (AF) is a major public health problem and one of the commonest supraventricular arrhythmias. At least 20% of all strokes are directly attributable to AF. However, its burden is not clear in sub-Saharan Africa, possibly due in part to undervaluation and non-recognition. In recent decades several new devices have been developed for the betterment of the accuracy and rates of AF detection, which also offers flexibility and feasibility.

**Objective:** The objective of the study is to identify the prevalence of known and Unknown AF in a high-risk patient population using a validated single-lead ECG.

**Methods and materials:** An institution-based cross-sectional study was conducted in 410 adults (≥55 years) recruited from Tikur Anbessa hospital sub-specialty outpatient clinics from July to October 2021. ECG tracing was conducted using (AliveCor) SL ECG device. A standardized questionnaire was designed to collect socio-demographic and clinical information using ODK Collect v2021. 3.0. we used logistic regression to determine the potential associated factors.

**Results:** The overall prevalence of AF in our study was 4.8%. It was strongly associated with age, diagnosis of heart failure, Substance use and high monthly income according to World Bank (WB) category. All the newly diagnosed patients were not on anticoagulation for stroke prevention.

**Conclusion and Recommendations:** Single timepoint screening in high-risk patients identified previously unknown AF in 2.1 %, out of which the majority were eligible for anticoagulation therapy. Identification of AF through targeted screening with up incoming novel technologies could reduce the stroke burden associated with undiagnosed AF.

## 1. Introduction

### 1.1. Background

Atrial Fibrillation (AF) is a major public health problem and one of the commonest supraventricular arrythmias.^1^ Early identification of atrial fibrillation (AF) is emerging as a priority in cardiology practice because of its association with substantial morbidity and mortality. AF increases cardiovascular disease risk, resulting in a five-fold increased stroke risk.^23^At least 20% of all strokes are directly related to AF, and in 20-45% of cases AF is first diagnosed at the time of stroke^4^. This value is likely an underestimate because a sizable proportion of cryptogenic stroke is due to undetected AF. ^5,6^ Patients with AF don’t only have a high risk of overt stroke they are also at an increased risk of a silent vascular brain lesion. Evidently, both Alzheimer’s and Vascular dementia were diagnosed in participants with no history of a clinical stroke and after accounting for possible stroke risk factors in a recent Korean study.^7,8^

Atrial fibrillation (AF) is an epidemic with significant health and economic impact. In 2010, there were around 33 million people worldwide estimated to have AF and this value is expected to double by 2050. ^9^ Its burden is not clear in sub-Saharan Africa, possibly due in part to under valuation and non-recognition.^10,11^There are only a hand full of population screening studies for AF across the African continent, and these studies have reported relatively low AF burden, together with a very significant heart failure and stroke burden. This shows more robust evidence from large scale screening studies is not yet available. ^12^

Commonly used AF screening strategies are opportunistic or systematic screening of individuals with risk factors particularly people, above the age of >_65 years or with other risk factors suggestive of increased stroke risk. These groups are usually screened by single-point or repeated 30-s ECG recording over a period of 2 weeks. Multiple new devices have been developed to improve the accuracy and rates of AF detection, which also offer flexibility and feasibility.^13^One such device is the AliveCor SL ECG device. However, limited data exist on their use for AF screening in low-resource countries even though it is approved by the US Food and Drug Administration (FDA) for automatic classification of 30-secondsingle-lead ECG tracing as normal or possible AF. ^14^

“An ounce of prevention is worth a pound of cure” a famous quote by Benjamin Franklin emphasizing the need for screening. The World Health Organization lists different conditions that should be fulfilled to justify mass screening.^15^ AF meets all these criteria. Screening for malignancies conditions is recommended even though the findings might not impact the patients’ life expectancy, but in the case of AF screening has an instantaneous indication for OAC treatment in those individuals with high CHA2DS2-VASC score.

Therefore, the aim of this study is to determine the overall prevalence of known and new AF, identifiable by single time-point screening using single lead ECG, examine their CHA2DS2-VASC score and eligibility for OAC in those with previously missed AF and introduce the use SL ECG device in low resource setup.

### 1.2 Statement of The Problem

Atrial fibrillation has become a major public health issue of epidemic proportion. Although many patients with AF present with palpitations, there is likelihood that their initial manifestation might be a debilitating stroke or death. Diagnosing AF before symptoms manifest could lead to initiation of appropriate effective therapy. Data in the prevalence of AF are scarce in the sub-Saharan African population. Utilization of newer smart ECG devices could be an effective mechanism to assess prevalence and explore the applicability of the devices to cardiology practice in Ethiopia.

### 1.3 Significance of The Study

This study will provide data on the prevalence of AF in asymptomatic patients and identify a missed opportunity. Since early and timely detection of AF will help prevent stroke

## 2. Literature Review

### 2.1 Introduction

AF is characterized as an irregular, disorganized and rapid atrial activation with loss of atrial contraction causing an irregular ventricular rate. This response is determined by AV nodal conduction. Atrial Fibrillation (AF) is a significant public health problem and one of the commonest supraventricular arrythmias. ^1^

### 2.1 Prevalence of AF

Atrial Fibrillation burden has been on the rise since the 1990s with a doubling of cases in the last 3 decades, ^16^ Current world prevalence is about 1–3% in the general population but increases with increasing age (from 9% in those above the age of 65 to 17% in those above 80 years old), presence of co morbidities, male sex, ethnicity, region, and screening method used. ^17–21 22^According to the Global Burden of Disease study in 2010, the prevalence of AF (age-adjusted, per 100 000 population) was 659.8 (95% UI, 511.0–850.4) for men and 438.1 (95% UI, 340.2– 561.0) for women in Sub Saharan Africa. ^19^ However, the GBD study also pointed out the scarcity of data in Sub-Saharan Africa and recommended targeted population studies for a better estimate. ^23^ In addition this part of the world is undergoing a major epidemiological transition with adoption of western lifestyle and suffering the consequences that come with it. These are hypertension, dyslipidemia, diabetes, and obesity all risk factors for cardiovascular diseases. There is an even larger deficiency of evidence in Ethiopia. A study done in TikurAnbesa Specialized Hospital (TASH) reveals prevalence of AF in RHD was (46.8%), a high prevalence of cardio embolic event (9.2%). ^24^ Another Ethiopian study evaluating the overall prevalence of AF in a community-based cross-sectional study community done in Jimma town was alarmingly high at 4.3% compared to the 0.5% from the GBD 2010 study.^25^ To our knowledge this previous study is the only community survey done in the country. We couldn’t find any data on the prevalence of non-rheumatic AF in the Ethiopian context.

### 2.3 Complications of AF

Many patients with AF manifest symptoms leading to detection of the arrythmia and appropriate management. However, a significant number of patients can present with debilitating stroke more so than ischemic stroke secondary to arterial disease.^26^ Detecting asymptomatic AF could enable the initiation of appropriate therapy, including oral anticoagulants (OACs) and prevent stroke and related complications. The other benefit is potential initiation of risk-factor modifiers.^13,18,27^Oral anticoagulation is estimated to reduce stroke risk for people with AF by about 65% compared to placebo, ^3,28^ but the real-world evidence on the use of anticoagulation in deserving patients is low in one study in Europe only 29.5% were receiving oral anticoagulants. ^22^

### 2.4 AF Screening

Currently opportunistic pulse palpation together with 12 lead ECG screening is practiced as a strategy to increase timely AF detection rate in high-risk patients. This is mainly based on the 60% improvement in AF detection in the landmark SAFE trial. ^29^ Based on this the European society of cardiology (ESC) recommends opportunistic screening for those above the age of 65. Citing the above evidence, the National Institute for Health and Care Excellence (NICE) also recommends screening but has a more stringent criteria where screening is recommended for patients with symptoms suggestive of AF. ^13,30^

However, some international recommendations against screening exist. These are underpinned by three major explanations. First, the cost implications and uncertainty over the benefits of a systematic screening program compared to usual care^31^ like the US Preventive Services Task Force citing a metanalysis which 1-time systematic screening with ECG did not detect more cases than opportunistic screening with pulse palpation.^32^Second argument is based on the potential harms of screening with ECG like misinterpretation of ECGs strips by primary care physicians, resulting in unnecessary treatments for individuals without atrialfibrillation.^33^Third, argument made by authors is that under treatment of known atrial fibrillation has to be prioritized. This is based on evidence that indicate low rate of appropriate therapy in such patients. For instance, a Swedish study revealed that only 57% of patients were on OAC even though they already had an established AF diagnosis. ^9^

Multiple new devices have been developed to improve the accuracy and rates of AF detection. These new technologies produce rapid results and are deemed to be cost effective compared to prior methods like pulse checking and 12-lead ECG for AF screening. ^34,35^

Different studies and systematic reviews using one single ECG recording found new AF in 1.4% of subjects >65 years which is high enough to support the use of single time-point Screening. ^36^ Interestingly the stroke stop study found 0.5% new AF via the initial ECG recording but with intermittent systematic screening detection of new AF increased 4-fold to 3% of the screened population. ^37^This shows us that most paroxysmal AF can be missed using single time ECG. Which strengthens the argument for screening. While many devices have been developed for the detection of AF, the most appropriate approach to deploy depends on various factors such as underlying AF, stroke risk, convenience, and cost. ^38^

In general, experts have reviewed major knowledge gaps and identified critical research priorities one of which is role of opportunistic screening in AF. ^24^ As mentioned previously, the therapeutic and prognostic implications of screening AF are still debatable. Moreover, uniform consensus does not exist whether one approach is superior to the other. Therefore, during the National Heart, Lung, and Blood Institute’s virtual workshop which convened to identify key research priorities in the field flagged AF screening as one of the most important research priorities in 2021.^39^

## 3. OBJECTIVE

### 3.1 General Objective

The objective of the study Is to determine the prevalence of AF in a high-risk patient population using a validated single lead ECG

### 3.2 Specific Objectives

- To assess baseline characteristics of high-risk patients presenting to the OPD.
- Prevalen of known and unknown AF in high-risk patients attending outpatient clinics
- To assess the association between the CHA₂DS₂-VASc score and associated values with the occurrence of AF
- Examine thromboembolism risk and eligibility for treatment in those with previously undiagnosed AF.

## 4. METHODOLOGY

### 4.1. Study Setting

#### 4.1.1. Study Area

The study was conducted at TASH, the biggest tertiary health care center in Addis Ababa Ethiopia. The hospital provides specialized medical care to patients across the country who are referred for various specialty care programmes. The hospital also serves as the main medical training center for undergraduate, specialty training and fellowship trainings. The cardiac clinic caters close to 1500 patients per month, the outpatient service provides Echocardiography and 12 lead ECG services as well. All new cardiovascular patients are evaluated by a cardiologist.

#### 4.1.2. Study Period

The study was conducted in Addis Ababa Tikur Anbessa hospital (TASH) Cardiac sub-specialty outpatient clinics from July to October 2021.

### 4.2. Study Design

An institution based cross sectional study was conducted to achieve the aims of this study.

### 4.3. Source and Study population

All patients attending internal medicine cardiac, renal, and diabetic specialty clinics for the follow up of different illnesses during the study period.

### 4.4. Eligibility Criteria

#### 4.4.1. Inclusion Criteria

All adult patients above the age of 55 attending selected internal medicine (Diabetes, Renal and cardiac) specialty clinics were included.

#### 4.4.2. Exclusion Criteria

Patients who had an established diagnosis of Rheumatic heart disease and those that refused to give informed consent were excluded.

### 4.5 Sample size Determination and Sampling Technique

Formula for single population Proportion

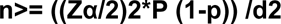

Where, p= Expected population proportion of the event: 4.3%

Q= 1-p

d= degree of absolute precision/ error tolerated (0.02)

Z α/2= Standard normal variable at 95% Confidence level (1.96)

A sample of 395 was calculated using a single population proportion formula assuming AF prevalence of 4.3 based on the Jimma community based cross sectional study 95% confidence level, 2 % margin of error.^25^

Simple random sampling was used to determine participation in the study.

### 4.6. Study Variables

#### 4.6.1. Outcome Variable

For this study the main dependent variable was the detection of AF

#### 4.6.2. Explanatory Variables

Independent/Explanatory Variables were selected based on extensive literature review and evidence-based assumption concerning their potential impact on AF.

### 4.7. Operational Definitions

Definite diagnosis of AF in screen-positive cases is established only after the physician reviews the single-lead ECG recording of >_30 s or 12-lead ECG and confirms that it shows AF.

### 4.8. Data Collection Procedures

#### 4.8.1. Data Collection Instruments

Clinical and laboratory profiles of participants were obtained by reviewing digital records of patients using data extraction tool. Structure questionnaire was prepared to collect socio demographic data from patients using the WHO STEPS Instrument for Chronic Disease Risk Factor Surveillance.^42^ The questionnaire was prepared in English and translated into Amharic and re-translated back to check its consistency (<u>appendix VI</u>). A pretest was conducted before the actual data collection begun.

Patients underwent AF screening using the AliveCor Heart Monitor^1^. This SL-ECG consists of a pair of electrodes and produces results corresponding to the lead I in the 12-lead ECG.^43^ For all patient’s fingers of each hand are placed on each electrode and the ECG recorded for about 30 seconds which instantly transmits tracings to a device with the KardiaMobile Application. We used a Samsung device (version5.18.2) that produces automated reports of ECG readings as ‘Normal’, ‘AF’, or ‘Unclassified’. A 12-lead ECG was performed for the subjects identified as having AF on the SL ECG device, and all 12-lead ECGs were reviewed by the first author of this manuscript.

**Figure 1:**
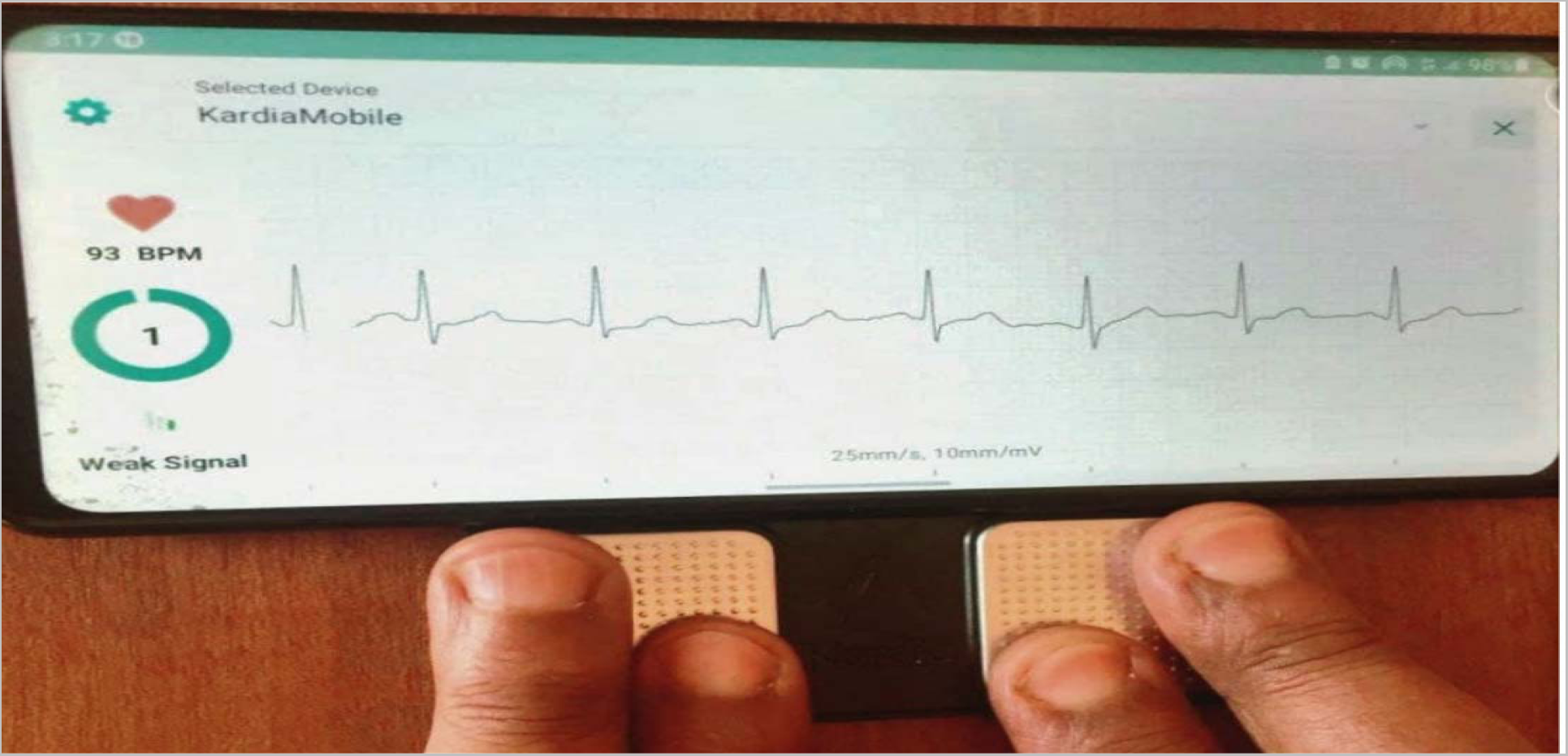
Patient undergoing SL ECG examination.

Participant’s anthropometry was assessed during the study. Weight was taken using standard beam balance and scale. Height was measured using standard measuring scale

Data was collected using ODK Collect v2021. 3.0 and stored on the Kobo Toolbox server. Four medical interns were recruited and given adequate familiarization training before the commencement of data collection.

#### 4.8.2. Data quality and management

The data was checked for completeness and consistency at the end of each data collection day and continuous supervision and monitoring took place during the whole duration. The data was explored and cleaned. We assumed missingness of values to be completely at random (CRD) and hence we opted for complete case detection. Data cleaning processes were validated with the data collectors to confirm accuracy. Outliers were assessed and counter checked with patient files to double check entry and considered. String variables were uniformized to facilitate analysis. Some continuous variables (Age, BMI, estimated GFR) were categorized according to universally accepted definitions. Prior to bivariate analysis, relevant categorical variables are restructured to produce results easily adaptable to clinical settings.

#### 4.8.3 Data Analysis

Data was analyzed using Stata version 16.1 (StataCorp LP, College Station, TX, USA). Test statistics with p-values <0.05 were considered significant and CIs were calculated for a 95% level of confidence.

The distribution of each relevant variable was explored using graphical methods, where uncertainty exists this was substantiated by the D’Agostino’s K^2^ test. Essential sociodemographic and clinical characteristics were presented using standard descriptive statistics (counts, proportions, means, standard deviations (SD), medians, interquartile ranges (IQR)).

A bivariate analysis was performed to analyze associations between AF and potential predictor variables appropriate test statistics were used were relevant (see Appendix IV). A multivariable logistic regression was conducted to evaluate the compounded effects of associated factors and adjust for confounders. Potential independent variables were identified a priori from published literature. Model diagnostic tests were conducted to ensure the fitted logistic regression estimated the outcome to a reasonable extent. Hence, Hosmer-Lemeshow-Test (HLT) and post model ROC (Receiving Operating Characteristic) curve was plotted.

### 4.9. Ethical consideration

Ethical clearance was obtained from the AAU college of health sciences Internal Medicine Department Ethical Review Board. The safety and privacy of subjects was protected by anonymizing identified information during data collection and analysis. Informed written consent was obtained from participants of the study (Appendix II).

### 4.10. Dissemination of the results

The final output of this study will be submitted to Addis Ababa University, College of health sciences. It will be presented at different seminars, journals, and workshops. Finally, efforts will be made to publish in peer-reviewed journals.

## 5. Results

### 5.1 Descriptive Analysis

#### 5.1.1 Sociodemographic characteristics

A total of 410 patients attending three internal medicine specialty clinics (Diabetes, Renal and Cardiac) took part in screening. 15 patients were excluded from the dataset because of previous history of valvular heart disease and incomplete history of comorbidities. A total of 395 patients were included in the final analysis of the study. The comparison of baseline characteristics of the patients is presented in the (table 3) below. More than half of the study participants were female (n=207, (52.42%)). The median age is 65.00 (IQR 58.00-72.00) and most of the participants are under the age of 65. (N=236 (59.75%)). More than half of the participants (48.1%) were from Oromia, (30.13%) from Amhara and about 13% from SNNPR with a small fraction from other regions in Ethiopia.

Out of all participants, 308 (77.97%) were married and 44 (11.14%) were widowed the remaining ones were either single or divorced. Most participants have completed primary Education 179(42.32%) and have average income per month (n=106, 26.84%) based on the world bank’s classification. Occupation wise majority of our participants were housewife and employed by the government. (n=110, 27,85% and 70, 17.72% respectively).

#### 5.1.2 Clinical characteristics of patients

The most frequent diagnosis among participants was Type 2 DM (n=229, 57.97%), followed by hypertension (n=218, 55.19%) and Heart failure (n=80, 20.25%). All patients with heart failure have an echocardiography result showing the cause is not a valvular heart disease. Concerning anthropometric indices, the mean BMI was (24.93 +- 3.85), with more than half of the participants (55.44%) being either overweight (n=159, 40.25) or obese (n=60, 15.19%). Only about 169 (42.78%) of our participants had their Lipid profile checked (Registered) in the past 6 months. For those that had registered the median LDL value was 105 (83-126). Complete clinical characteristics of patients and the comparison of values in those with and without Atrial Fibrillation is presented in table 3 below.

**Table 1.** Sample size calculation.

| Objective | Formula | Assumption | Sample size |
| --- | --- | --- | --- |
| Screening of AF in high-risk patients attending outpatient clinics | $n \geq ((Z\alpha/2)^2 * P (1-p)) / d^2$ | P: 4.3%<br>D: 0.02<br>95% Confidence level (1.96) | 395 |
| Prevalence of AF in high-risk patients attending outpatient clinics | $n \geq ((Z\alpha/2)^2 * P (1-p)) / d^2$ | P: 7.2%<br>D: 0.03<br>95% Confidence level (1.96)<br>P was taken from a UK study | 285 |
| To assess the association between the CHADS2VAS score and associated values with the occurrence of AF | $n \geq ((Z\alpha/2)^2 * P (1-p)) / d^2$ | P: 20%<br>D: 0.05<br>95% Confidence level (1.96)<br>P was taken from the Rotterdam study <sup>(40)</sup> | 245 |
| Examine thromboembolism risk and eligibility for treatment in those with previously undiagnosed high-risk patients with AF | $n \geq ((Z\alpha/2)^2 * P (1-p)) / d^2$ | P: 30%<br>D: 0.05<br>95% Confidence level (1.96)<br>P was taken from the Framingham study <sup>41</sup> | 322 |

**Table 2.** Study Variables.

|  |
| --- |
| **Patient Socio-Demographic Characteristics** |
| Age<br>Gender<br>Income category (based on WB category)<br>Level of education<br>Marital status<br>Ethnicity |
| **Anthropometric Measurements** |
| BMI |
| **Clinical and Laboratory Characteristics** |
| Diagnosis of Diabetes<br>Diagnosis of Hypertension<br>Diagnosis of Heart Failure<br>Diagnosis of Anemia<br>Diagnosis of COPD<br>Diagnosis of IHD<br>LDL<br>Estimated GFR |
| **Behavioral Factors** |
| Smocking Status<br>Alcohol consumption<br>Khat Chewing |
| **Treatment Characteristics** |
| Oral Anticoagulation use<br>Aspirin Use |

**Table 3.** Comparison of baseline characteristics.

| Variable | New AF (N=8) | Known AF (N=11) | No AF (N=376) |
| --- | --- | --- | --- |
| **Sex** |  |  |  |
| Male, n (%) | 5 (62.50) | 5(45.45) | 178(47.34) |
| Female | 3(37.50) | 6(54.55) | 198(52.66) |
| **Age Categories(years), n(%)** |  |  |  |
| 55-65 | 3(37.50) | 4(36.36) | 230(61.17) |
| 65-75 | 1(12.50) | 7(63.64) | 99(26.33) |
| >75 | 4(50.00) | 0(0) | 47(12.50) |
| **Marital Status, n (%)** |  |  |  |
| Single | 1(12.50) | 0(0) | 28(7.45) |
| Married | 6(75.00) | 11(100) | 29(77.39) |
| Divorced | 0 | 0 | 14(3.72) |
| Widowed | 1(12.50) | 0 | 43(11.44) |
| **Ethnicity, n (%)** |  |  |  |
| Amhara | 3(37.50) | 4(36.36) | 112(29.79) |
| Gumuz | 0 | 0 | 1(0.27) |
| Oromo | 4(50.00) | 4(36.36) | 182(48.4) |
| SNNPR | 0 | 3(27.27) | 50(13.30) |
| Sidama | 0 | 0 | 4(1.06) |
| Somalia | 0 | 0 | 1(0.27) |
| Tigray | 1(12.5) | 0 | 26(6.91) |
| **Occupation, n (%)** |  |  |  |
| Housewife | 1(12.50) | 3(27.27) | 106(28.19) |
| Farmer | 2(25.00) | 2(18.18) | 21(5.59) |
| Civil Servant | 2(25.00) | 1(9.09) | 67(17.82) |
| Merchant | 0(0) | 1(9.09) | 18(4.79) |
| Others | 3(37.50) | 4(36.36) | 164(43.62) |
| **Income Status n, (%)** |  |  |  |
| Low Income | 7(87.50) | 6(54.55) | 259(68.88) |
| Average Income | 0 | 3(27.27) | 103(27.39) |
| High Income | 1(12.50) | 2(18.18) | 14(3.72) |
| **Education n, (%)** |  |  |  |
| Primary Education | 4(50.00) | 5(45.45) | 169(44.95) |
| Secondary Education | 3(37.50) | 3(27.27) | 95(25.27) |
| Higher Education | 1(12.50) | 3(27.27) | 112(29.79) |
| **BMI (kg/m<sup>2</sup>) categorized, n (%)** |  |  |  |
| <18.5 | 0(0) | 0(0) | 14(3.72) |
| 18.5-24.9 | 4(50) | 5(45.45) | 153(40.69) |
| 25.0-29.9 | 4(50) | 5(45.45) | 150(39.89) |
| >30 | 0 | 1(9.09) | 59(15.69) |
| **Pulse Rate(bpm), mean** | 109.25(+/-33.59) | 105.90(+/-37.11) | 86.71(+/-86.71) |
| **Estimated GFR<sup>2</sup>, categorized, n (%)** |  |  |  |
| Group 1 (Normal > 90) | 5(62.50) | 7(63.64) | 317(84.31) |
| Group 1 (eGFR 60-90) | 2(25.00) | 3(27.27) | 31(8.24) |
| Group 2 (eGFR 30-60) | 1(12.50) | 1(9.09) | 18(4.79) |
| Group 3 (eGFR 29-30) | - | - | 9(2.39) |
| Group4 (eGFR <15) | - | - | 1(0.27) |
| **Medical History, n (%)** |  |  |  |
| Diabetes | 2(25.00) | 10(90.91) | 217(57.71) |
| IHD | 2(25.00) | 1(9.09) | 27(7.23) |
| Heart failure | 1(12.50) | 6(54.55) | 73(19.41) |
| Asthma | 0(0) | 1(9.09) | 21(5.59) |
| COPD | 0(0) | 0(0) | 3(0.80) |
| Anemia | 1(12.50) | 1(9.09) | 23(6.12) |
| CKD | 2(21.05) | 2(18.18) | 57(15.16) |
| Hypertension | 3(37.50) | 9(81.82) | 206(54.79) |
| Hyperthyroidism | 0(0) | 0(0) | 19(5.05) |
| **CHA<sub>2</sub>DS<sub>2</sub>-VAsC n (%)** |  |  |  |
| Score <2 | 1(12.5) | 1(9.09) | 81(21.54) |
| Score ≥ 2 | 7(87.5) | 10(90.91) | 295(78.46) |
| **Substance Use n (%)** |  |  |  |
| Alcohol | 3(37.50) | 1(9.09) | 31(8.24) |
| Smoking | 2(25.00) | 0(0) | 13(3.46) |
| Chat Chewing | 0(0) | 0(0) | 18(4.79) |

<sup>2</sup> GFR is estimated and categorized using the recommendations by CKD-EPI equation.

| Variable | New AF (N=8) | Known AF (N=11) | No AF (N=376) |
| --- | --- | --- | --- |
| **LDL (mg/dl), n(%)** |  |  |  |
| LDL < 100 | 3(37.5) | 3(27.27) |  |
| LDL ≥ 100 | 5(62.5) | 8(72.73) |  |
| **Treatment n, (%)** |  |  |  |
| Aspirin | 0(0) | 2(18.18) | 117 |
| Warfarin | 0(0) | 9(81.82) | 16 |

### 5.2 Prevalence of AF

Of the initial single-lead ECG tracings obtained from 395 participants, the KardiaMobile algorithm was able to provide a rhythm decision in 86% (340/395) of screened patients the rest of the outputs indicated unclassified (19/395,4.81%), unreadable (36/395, 9.11%) recordings. Samples of ECG Kardia output are presented in the appendix (<u>appendix VI</u>). A repeat tracing was obtained in 49 (89%) of the participants who did not have an initial rhythm decision. Of those participants without repeat KardiaMobile tracings, 100% (19/55) had an initial result of unclassified, which the screening team deemed as sinus tachycardia (>100 bpm) and interpreted as normal rhythm not requiring a repeat tracing,

AF was detected in 22 patients (5.57%) using the single lead ECG. Manual confirmation of the tracings indicated that 3 of the patients were erroneously classified AF by automated reading and when a confirmatory 12 Lead ECG was obtained these patients turned out to have sinus arrythmia, RBBB and sinus rhythm. All the 22 patients received 12 lead ECG to confirm the diagnosis of AF. Out of these A previous diagnosis of Non valvular AF was present in 11 patients (2.78%), the majority of whom were men (n=10, 52.63%)). The overall prevalence of New AF detected by SL-ECG or known AF was 4.81% (19/395) with similarly a higher prevalence in men (n=10, 52.63%) than women.

**Figure 2.**
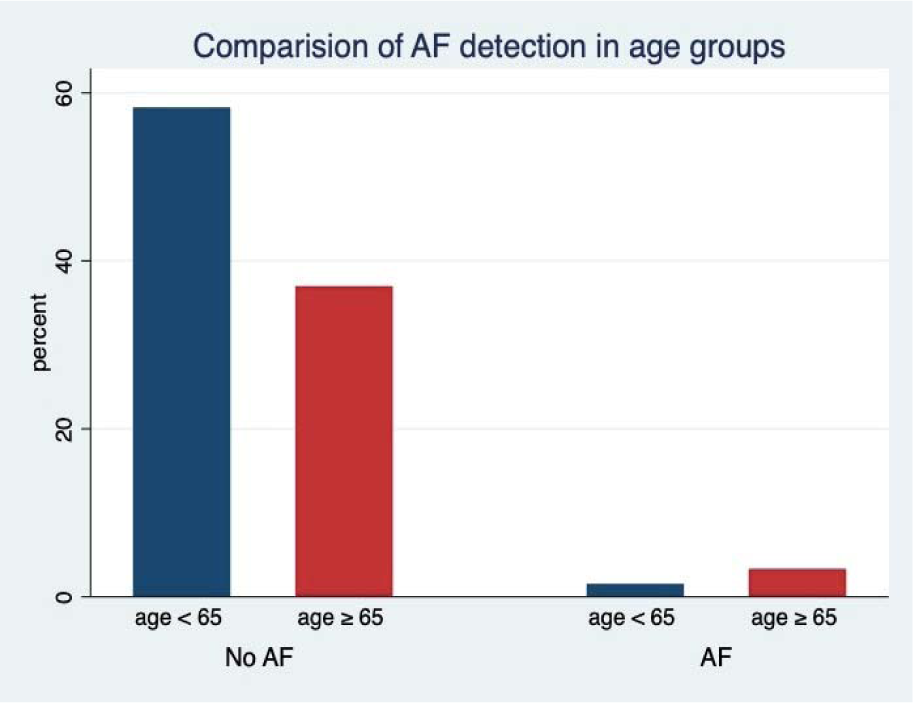
Comparison of AF detection among age groups.

### 5.3 Ability to detect newly diagnosed AF by screening and PPV of SL-ECG

Of the AF detected from 395 participants, new AF was detected in 8 patients (2.03%), known AF was detected in 11 patients. In total the prevalence of AF was determined to be 4.8% in the study population. There was a 42.1% (8/19) increase in the prevalence rate of AF with this screening programme. The PPV of the SL ECG calculated in our study is 86.3%.

### 5.4 Factors Associated With AF

Patients’ sociodemographic and medical characteristics were analyzed to assess their effect on the development of AF. Eleven variables were selected for initial univariate analysis. Of this age category > 65 (COR (95%CI), P value) = 3.41 (1.27-9.18) (0.015), Any substance use, and high-income category were found to have strong evidence of independent association with AF.

In the multivariable analysis, Age, the diagnosis of heart failure, and substance use showed strong evidence of association. Complete results of the multivariable analysis are presented in the (table 4) Accordingly age was the strongest predictor for developing AF (AOR 2.08(1.11-3.89)) (*p=*0.02). We conducted post model estimation tests to assess the fitness of the logistic model. Hosmer Lemishow Test indicates that the specified model fits reasonably well Additionally, the post model ROC curve indicates that the area under the curve is approximately 0.79 indicates acceptable discrimination for the model.

**Table 4.** Bivariate logistic regression analysis of selected variables.

| Variable | COR (95% CI) | P-value |
| --- | --- | --- |
| **Sex** |  |  |
| Male | 1.24(0.49-3.11) | 0.653 |
| **Age** |  |  |
| 55-65 | Reference |  |
| 65-75 | 2.66(0.93-7.52) | **0.06** |
| >75 | 2.80(0.79-9.94) | 0.11 |
| **Marital Status** |  |  |
| Divorced | Reference |  |
| Married | 2.51 (0.33 - 19.36) | 0.38 |
| Single | 1.54 (.092-25.57) | 0.77 |
| **BMI (kg/m<sup>2</sup>)** |  |  |
| <18.5 | (No value) |  |
| 18.5-24.9 | Reference |  |
| 25-29.9 | 1.02(0.39-2.64) | 0.96 |
| >30 | 0.29(0.36-2.32) | 0.26 |
| **Gfr** |  |  |
| Group 1 (eGFR 60-90) | 4.26(1.40-12.88) | 0.01 |
| Group 2 (eGFR 30-60) | 2.93(0.61-14.11) | 0.17 |
| Group 3 (eGFR 29-30) | 1 |  |
| Group4 (eGFR <15) | 1 |  |
| **DM** | 1.26(0.48-3.26) | 0.64 |
| **HTN** | 1.41 (0.54-3.67) | 0.48 |
| **Substance Use (any)** | 3.37(1.15-9.91) | **0.03** |
| LDL < 100 | Reference |  |
| LDL ≥ 100 | 0.44(0.16-1.19) | 0.11 |
| **Heart Failure** | 2.42(0.92- 6.37) | **0.073** |
| **CHA<sub>2</sub>DS<sub>2</sub>-VASc Score ≥ 2** | 2.33(0.52-10.31) | 0.26 |
| **Income (As compared to low income)** | Reference |  |
| Average Income | 1.72(0.48-6.17) | 0.40 |
| High Income | 7.35 (1.35-40.07) | **0.02** |

**Table 5.**
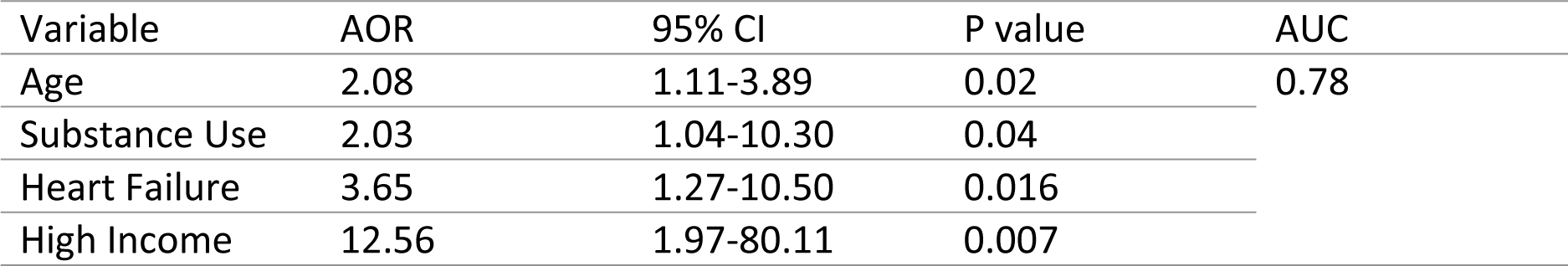
Multivariate regression output of selected variables.

| Variable | AOR | 95% CI | P value | AUC |
| --- | --- | --- | --- | --- |
| Age | 2.08 | 1.11-3.89 | 0.02 | 0.78 |
| Substance Use | 2.03 | 1.04-10.30 | 0.04 |  |
| Heart Failure | 3.65 | 1.27-10.50 | 0.016 |  |
| High Income | 12.56 | 1.97-80.11 | 0.007 |  |

**Figure 3.**
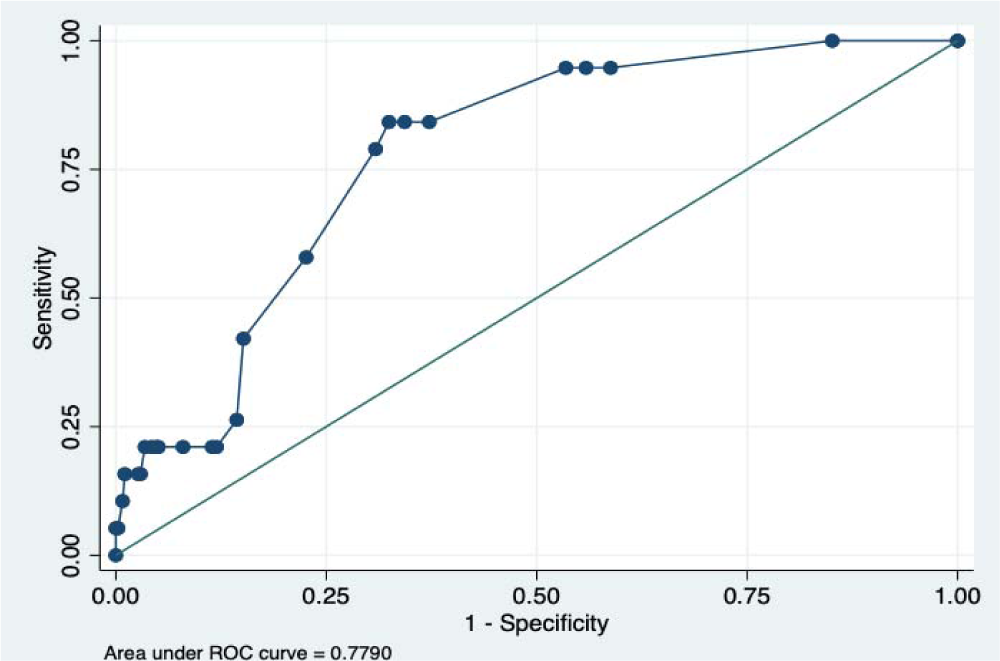
Post logistic regression ROC Curve.

Hence, we can assume the above stated logistic regression model can predict the occurrence of AF reasonably well.

### 5.4 Stroke Risk Stratification

Risk stratification was done using CHA2DS2-VASc Score. Accordingly, more than two third of the patients (n= 312, 78.99%) had a score of two or more. All the patients who had already known AF had a CHA2DS2-VASc score greater or equal to two, and 9(81.81%) are on warfarin and the rest of them are on aspirin. However, none of the patients who were newly diagnosed were on anticoagulants. Hence the prevalence of untreated AF among all the participants was 2.53%.

**Figure 4.**
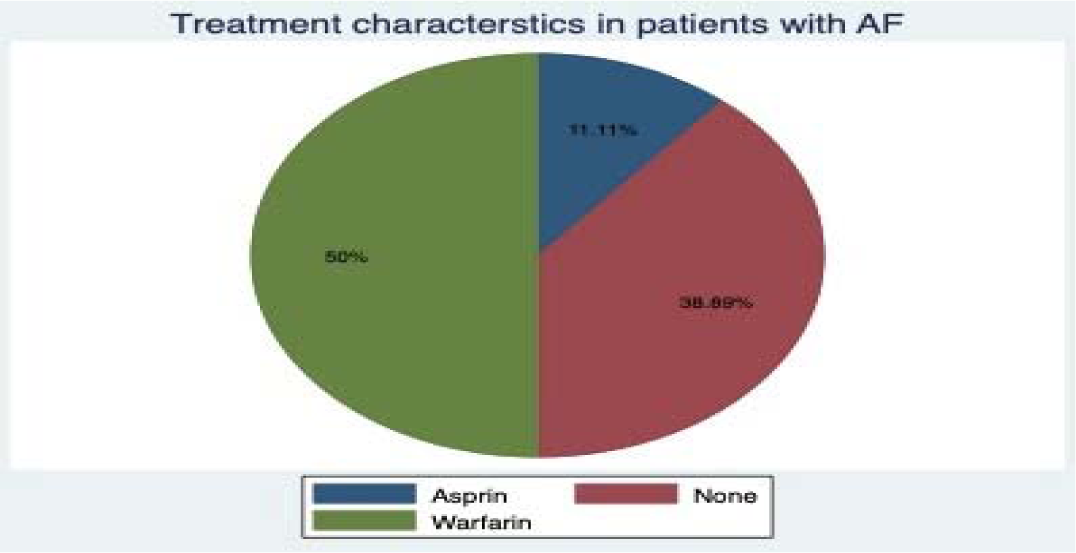
Treatment characteristics in patients with AF.

## 6. Discussion

To the best of our knowledge this is the first systematic non valvular AF screening study in high-risk patients using Single lead ECG recordings in the country. AF was confirmed in 19 patients (4.81%) using the single lead ECG. Out of these A previous diagnosis of Non valvular AF was present in 11 patients (2.78%), and New AF was detected in 8 patients (2.03%).

For a disease to be suitable for screening according to WHO one of the most important criteria is a screening test should be suitable and acceptable to the population^44^. Recent advances in wireless technology have enabled ECG screening for cardiac arrhythmias without the use of 12-lead ECG machines. In our current study we used a wireless smartphone-based device which is a simple, small and portable device, even though 12 lead ECG is the gold standard, it needs a private room, trained staff and frequent cleaning of the cables especially in the covid era which are luxuries scares in low resource setup like ours.^45 46^

In our study we found 8 patients (2.03%) with new AF diagnosis which is comparable to the metanalysis of screening studies which found 1.4% new AF in patients above the age of 65^47^. We think this number is underestimated since one time screening can miss cases of paroxysmal AF, as shown in the stroke stop study where AF diagnosis rose from 0.5% from initial electrocardiographic screening to (3.0%; 95% CI, 2.7–3.5) during 2 weeks of twice-daily electrocardiographic recording.^43^ The overall prevalence of AF was 4.8% in our study which is comparable to a study done in west Ethiopia where 4.3% prevalence of AF was documented.^25^ It is also comparable to studies done in US AND European populations; a study done from large California health maintenance organization reported an overall prevalence of 5.3% where the prevalence among whites, blacks, Asians, and Hispanics was 8.0%, 3.8%, 3.9%, and 3.6%, respectively^48^ compared to the studies above we have excluded patients with rheumatic heart disease. We have found the positive predictive value of our test to be 86.3% showing it has a low false positive rate compared to other studies which have reported different PPV values ranging from 55%-100%.^47^

Africa is going an epidemiologic shift fueled by urbanization, increased consumption of western diet and lifestyle. According to previous studies, up to two-thirds of cases of AF in Ethiopian patients could be attributed to RHD^49–51^ . we need to challenge this assumption as nonvalvular AF is associated with the afore-mentioned risk factors. Many studies have identified age, male gender, diabetes, hypertension, and heart failure to be risk factors for non-valvular AF. In our study, we have identified age, heart failure, substance use and high-income status, as independent predictors of AF. In our study there was no difference between men and women regarding risk of AF, but male sex has been ascribed as a risk in the Framingham study. ^41^

Age is one of the strongest predictors of AF in our study with each advancing decade the risk of AF doubles which is similar compared to the Framingham and other epidemiological studies.^41,52^ cigarette smoking is positively correlated with AF but just short of significance although when its was combined with alcohol use it became a strong predictor of AF.

Other cardiovascular risk factors like hypertension and diabetes were not significantly associated even though they are positively correlated. The reason could be most of our patient population was from diabetic clinic.

Screening a particular disease is justified if there is an adequate and effective treatment once the disease is identified. In the case of AF OAC is an effective way of preventing stroke it can reduce stroke by approximately 60% and death by approximately 25% compared with no antithrombotic treatment ^53^. In our study 9 out of the 11 (2.3%) known AF and none of the New AF patients were on anticoagulation making the prevalence of untreated AF 2.53%. This is a missed opportunity, and the gap needs to be filled by implementing screening procedures equipped with easy-to-use technological advances.

**Figure 5.**
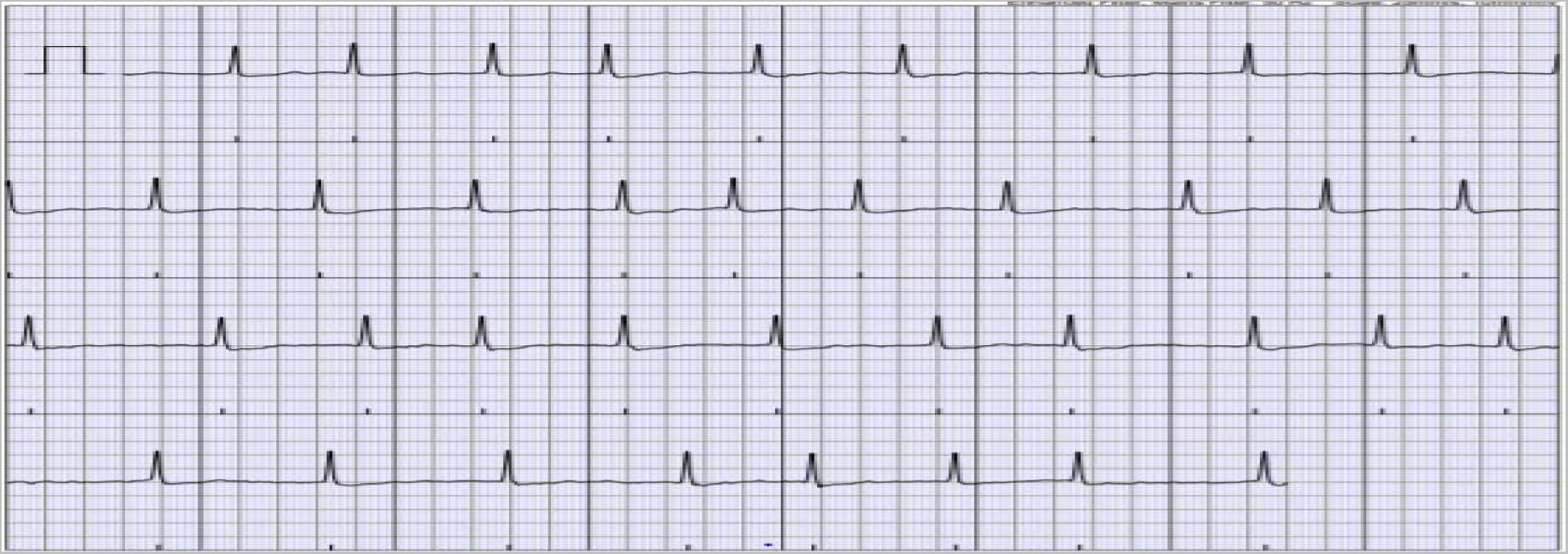
AF by SL ECG Device.

## 7. Recommendations, Conclusion

Single timepoint screening in high-risk patients identified previously unknown AF in 2.1 % of the study population, this is significant because most individuals diagnosed with AF have higher stroke risk. Hence, thombophylaxis is strongly indicated. Based on the results of this study we argue that targeted screening to detect new AF could contribute to the reduction of stroke and other thromboembolic events. This study has also demonstrated that novel technologies such as Alive core SL device, have simplified the diagnosis of AF and can facilitate implementation of systematic targeted screening in low resource setups.

We recommended screening of atrial fibrillation in selected high-risk patients with simple, easy to use devices like the one used in the study starting from our high-risk clinics. We also recommend this device to be used for larger studies to do population screening so that the true AF prevalence in Ethiopian population can be estimated.

## 8. Limitation of The Study

The study utilized online tools to collect and ensure quality of data. We also utilized a relatively novel technology of screening AF. However, this study has several limitations, first it was not a large population scale screening and hence generalizability of the results to other settings needs a lot of caution and can be challenging. Second cost effectiveness analysis was not done, third 12 lead ECG was not performed for every study participant designated as sinus rhythm by the SL ECG. Subsequently we were not able to do sensitivity and specificity of the SL device compared to the gold standard even though the device has been validated elsewhere.

## ABBREVIATIONS AND ACRONYMS

AAU: Addis Ababa University
AF: Atrial Fibrillation
CHS: College of Health science
ECG: Electrocardiography
ESC: European society of cardiology
ETB: Ethiopian Birr
GFR: Glomerular Filtration Rate
IHD: Ischemic Heart Disease
LDL: Low-density lipoproteins
NICE: National Institute for Health and Care Excellence
OAC: Oral Anticoagulation
SL: Single Lead
TASH: Tikur Anbessa Specialized Hospital
WB: World Bank
WHO: World Health Organization

## Data Availability

All data is available in the manuscript

## Footnotes

1 AliveCor SL ECG device is <u>Cleared by the United States Food and Drug Administration</u>, CE marked and clinically validated.

